# Statin use and risk of amyotrophic lateral sclerosis: An active-comparator, new-user cohort study

**DOI:** 10.64898/2025.12.23.25342802

**Authors:** Yusuke Okada, Satoru Morimoto, Shinichi Takahashi, Hideyuki Okano, Hisashi Urushihara

## Abstract

**Importance:** Elevated circulating low-density lipoprotein (LDL) cholesterol is associated with an increased risk of amyotrophic lateral sclerosis (ALS) onset. Previous studies have explored the relationship between statin use and ALS onset. However, findings have been inconsistent, potentially due to methodological limitations, such as confounding by indication, and failure to account for baseline differences in LDL cholesterol levels.

**Objective:** To compare the risk of ALS onset between new users of statins and ezetimibe among patients with hypercholesterolemia.

**Design:** Active-comparator, new-user cohort study using inverse probability of treatment–weighted Cox proportional hazards models. The study period spanned April 2012 to February 2024.

**Setting:** Two administrative claims databases in Japan.

**Participants:** Patients with hypercholesterolemia who newly initiated statins or ezetimibe, and patients with dyslipidemia who newly initiated fibrates. All participants were required to have at least 365 days of baseline observation and no prior diagnosis of ALS.

**Exposure(s):** Statin use compared with ezetimibe use. Fibrate use was assessed for benchmarking.

**Main Outcome(s) and Measure(s):** The outcome was incident ALS, defined as a first definitive diagnosis of ALS. Hazard ratios (HRs) and 95% confidence intervals (CIs) were estimated to compare ALS risk between statins and ezetimibe.

**Results:** The study included 607,292 statin users (median [IQR] age, 61 [51–71] years; 51.4% male), 26,963 ezetimibe users (median [IQR] age, 59 [49–71] years; 50.1% male), and 114,871 fibrate users (median [IQR] age, 61 [51–71] years; 51.4% male). The incidence rate per 100,000 person-years of ALS was 6.8, 15.9, and 4.3, respectively. Statin use was associated with a lower hazard of ALS onset than ezetimibe use (adjusted HR [95% CI]: 0.42 [0.19–0.92]). The mean (SD) LDL cholesterol immediately prior to treatment initiation was 171.0 (28.6) mg/dL in the statin group and 162.8 (30.8) mg/dL in the ezetimibe group. After treatment, mean LDL cholesterol levels decreased to and stayed below 140 mg/dL in both groups.

**Conclusions and Relevance:** This study suggests that statins may lower the risk of ALS onset among patients with hypercholesterolemia. The mechanism underlying this association is not yet clear and may involve pathways beyond circulating LDL cholesterol reduction.

**Key points:** *Question:* Does statin use lower the risk of amyotrophic lateral sclerosis (ALS) onset among patients with hypercholesterolemia?

*Findings:* In this active-comparator, new-user cohort study of 607,292 statin users and 26,963 ezetimibe users with hypercholesterolemia, statin use was associated with a lower hazard of ALS onset compared with ezetimibe use. Prior to treatment initiation, mean LDL cholesterol levels were similar between statin and ezetimibe users.

*Meaning:* These findings suggest that statins may lower the risk of ALS among patients with hypercholesterolemia.

## Introduction

Amyotrophic lateral sclerosis (ALS) is a progressive and fatal neurodegenerative disease characterized by the selective loss of upper and lower motor neurons, resulting in progressive muscle weakness, atrophy, and ultimately respiratory failure.^1,2^ The disease remains incurable, and currently available therapies, such as riluzole and edaravone, offer only limited benefits in slowing disease progression or extending survival.^3,4^

Recent evidence suggests that abnormalities in lipid metabolism, particularly cholesterol dysregulation, may be involved in ALS pathogenesis, representing a new and promising avenue for therapeutic intervention.^5^ Several clinical studies reported that elevated circulating low-density lipoprotein (LDL) cholesterol is associated with an increased risk of ALS onset,^6–9^ and *in vitro* and *in vivo* experimental studies reported that excessive cholesterol biosynthesis in motor neurons elevated the neurotoxic cholesterol metabolite 24(S)-hydroxycholesterol.^10–12^

Sterol regulatory element-binding protein 2 (SREBP2) is a key regulator of cholesterol biosynthesis including the mevalonate pathway.^13^ Overexpression of SREBP2 in the spinal cord of normal mice increases cholesterol synthesis and induces ALS-like symptoms.^14^ As cholesterol cannot cross the blood–brain barrier (BBB), the relevance of cholesterol levels in the peripheral circulation and in the brain must be considered separately when investigating the pathogenesis of ALS.

Statins are widely used to treat dyslipidemia by suppressing LDL cholesterol synthesis, lowering the risk of cardiovascular disease consequences.^15^ Several studies have explored the association of statin use with ALS onset, however, the findings remain inconsistent, with the results ranging from no association,^16,17^ to an identified protective effect,^18,19^ to a potential increase in ALS risk.^20,21^ The causal contrasts used in prior studies may not have been appropriate for estimating the net effect of statins, because their comparison groups often included patients without lipid abnormalities. This potentially introduced heterogeneity in baseline lipid profiles, creating confounding factors that compromise comparison validity. In addition, statins differ in their lipophilicity, which may affect its permeability across BBB to central nervous system (CNS).^22^

Here, we applied an active-comparator, new-user design in patients with hypercholesterolemia to achieve a valid comparison and isolate the net effect of statins on ALS risk.

## Methods

### Data Source

We used two Japanese administrative claims databases: the JMDC claims database maintained by JMDC Inc. (Tokyo, Japan)^23^ and the DeSC database maintained by DeSC Healthcare, Inc. (Tokyo, Japan).^24^ The JMDC claims database includes claims data from the Corporate Health Insurance Society covering roughly 16 million insured persons from April 2005 through February 2024, whereas the DeSC database contains claims data from the Corporate Health Insurance Society, the National Health Insurance, and the Latter-Stage Elderly Health Care, representing approximately 12 million beneficiaries between April 2014 and November 2023.

The data period available for the study spanned April 1, 2014, to November 30, 2023 in the DeSC database and April 1, 2012, to February 29, 2024, in the JMDC claims database. The analysis dataset was constructed using both databases which had been irreversibly anonymized. In combining these two databases, we excluded the claims records from the Corporate Health Insurance Society in the DeSC database, to avoid potential duplicates of the same enrollee’s records as in the JMDC claims database. Individual consent to the collection and secondary use of existing personal health data was waived in accordance with the Japanese Privacy Protection Law. The study protocol was approved by the Ethics Committee for Research Involving Humans of Keio University Faculty of Pharmacy (No. 241016-2) and followed local ethical guidelines for biological and medical research involving human subjects.

### Study Design

We applied an active-comparator, new-user cohort design in patients with hypercholesterolemia to enhance comparison validity.^25^ A design diagram is shown in eFigure 1. The primary analysis compared statin initiators with ezetimibe initiators, as both drugs are used for the same indication but have different mechanisms of action. A secondary analysis compared lipophilic and hydrophilic statins.

### Study Population

The statin and ezetimibe cohort included patients who received a prescription of any statin or ezetimibe during the study period (eTable 1). The index date was defined as the first prescription date following a lookback period of 365 days with no prescription records of either drug. All cohort members were required to have at least one diagnosis record of hypercholesterolemia (ICD-10 code: E780) on or before the index date, and to have an observable period of at least 365 days before the index date in the database. We excluded the following patients; 1) who received multiple products of statin or ezetimibe on the index date, and 2) who had a diagnosis record of ALS on or before the index date, regardless of whether the diagnosis was flagged as suspected (eTable 2). Patients were grouped into either the statin or ezetimibe groups based on the drug prescribed on the index date. In the statin group, patient subgroups were further categorized based on lipophilicity (eTable 1).

A benchmark cohort consisting of the patients who received a prescription of a fibrate represented a non-hypercholesterolemia control (eTable 1). The fibrate cohort was required to have no prescription of statin or ezetimibe throughout the treatment with fibrate, and have at least one diagnosis record of dyslipidemia (ICD-10 code: E78) on or before the index date. Exclusion criteria were the same as those for the statin and ezetimibe cohorts.

For the purpose of calculating background incidence of ALS, we investigated all the subjects who were aged 40 years or older and continuously observable between July 2019 and December 2020 in the entire databases.

### Follow-up

The at-risk period for ALS began on the index date and ended 365 days after the last day of exposure to the study drug, as estimated by the last prescription. If the gap between the prescriptions was 100 days or less based on days of supply, the drug exposure was considered to be continuous. Follow-up commenced on the index date and was censored at the earliest of (i) completion of the predefined at-risk period, (ii) 365 days after initiation of, or add-on with, any non-index drug class, (iii) confirmed ALS onset, (iv) disenrollment from the health insurance system, or (v) termination of the database observation period.

### Outcome

Incident ALS was identified using the diagnosis records coded according to the Medical Information System Development (MEDIS) standard disease master. The MEDIS codes listed in the eTable 2 correspond to the ICD-10 G12.2 subclass for motor neuron disease. The case definition was developed with input from neurologists (S.M., S.T.) with expertise in ALS. ALS onset was defined as the date of the first recorded ALS diagnosis.

### Covariates

The covariates considered were age at the index date, sex, type of insurance, duration of hypercholesterolemia at the index date, use of lipid-lowering drugs other than the exposure drugs, antidiabetic drugs within the 30 days before and on the index date, and comorbidities such as hypertension, myocardial infarction, atherosclerosis, stroke, atrial fibrillation, heart failure, cancer, liver disease and chronic kidney disease within 12 months on or before the index date. Drug and disease codes are listed in eTable 3–4.

### Statistical Analysis

Summary statistics of baseline characteristics and the incidence rates of ALS were calculated by drug group: statins, statin subclass classified by lipophilicity, ezetimibe, and fibrates. Cumulative incidence curves were plotted by each drug group. In addition, the time series data of serum LDL cholesterol level measured at annual specific health check-up were plotted at 90-day intervals during the study period among a subset of drug group members who had at least one LDL cholesterol value was available.

A Cox proportional hazards model was used to estimate hazard ratios (HRs) and 95% confidence intervals (CIs) of ALS onset. Inverse probability of treatment weighting was used to balance baseline characteristics between groups. Propensity scores were estimated with logistic regression including all covariates above. Absolute standardized mean differences (aSMDs) < 0.10 was used for assessing covariate balance.^26^ We evaluated two comparisons in the Cox models, statin initiators versus ezetimibe initiators, and lipophilic versus hydrophilic statins. The incidence of ALS in the fibrate group was provided as the benchmark of dyslipidemia patients without elevation of LDL cholesterol.

We conducted the following sensitivity analyses to assess the robustness of main analysis results. We applied a stricter definition of the ALS outcome, which was a combination of the disease codes for ALS with a prescription for either riluzole or edaravone in the same month. Furthermore, we applied the different risk window lengths including 180 or 730-day post-treatment periods after treatment discontinuation or switching. We also applied a different treatment gap of 60 days when estimating the duration of continuous exposure.

All analyses were performed with SAS version 9.4 (SAS Institute Inc., Cary, NC, USA). A 2-sided P value of less than .05 was adopted as statistically significant.

### *In vitro* experimental study

To complement the cohort analysis, we conducted i*n vitro* experiments to assess neurotoxicity of ezetimibe, fenofibrate; details are described in eMethod 1.^27–29^ We did not test statins directly *in vitro* because the primary purpose of these experiments was to evaluate whether the comparator drugs (ezetimibe and fibrates) exhibited neurotoxicity that could bias the cohort comparison.

## Results

607,699 new users of statins, and 26,975 new users of ezetimibe, with a diagnosis of hypercholesterolemia were eligible for the study (Figure 1). Out of the new statin users, 282,493 were prescribed lipophilic statins, and 325,206 were hydrophilic statins. Separately, 114,943 were identified as new users of fibrates with a diagnosis of dyslipidemia.

**Figure 1.**
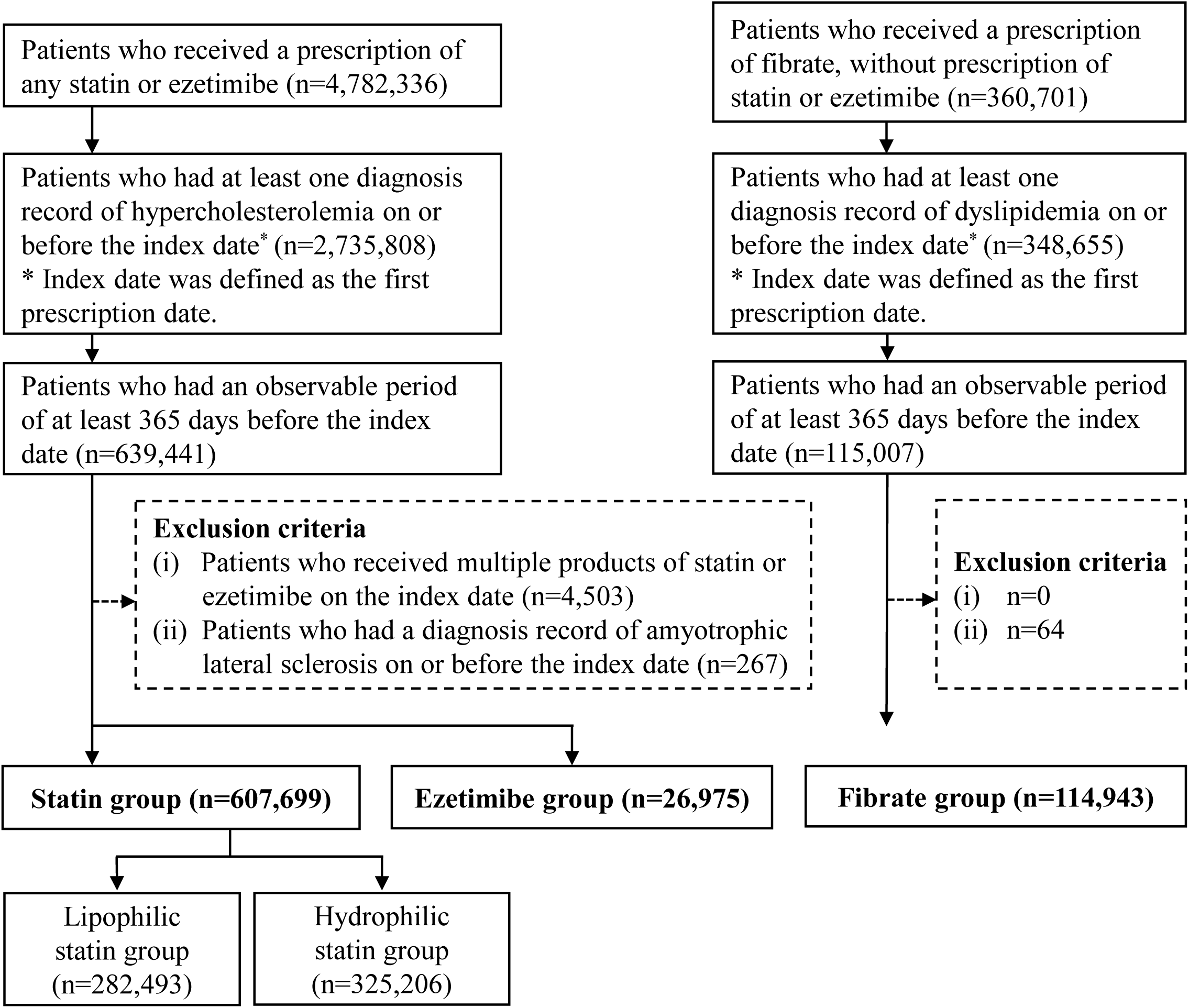
Patient selection flow. Flow chart of cohort construction. The figure shows the identification of eligible patients from the database, including baseline observation requirements, exclusion criteria, and the final numbers of statin initiators, ezetimibe initiators, and fibrate initiators included in the analyses.

Both the statin and ezetimibe groups had a median age around 60 years (statins: 61 [IQR, 51–71] years; ezetimibe: 59 [IQR, 49–71] years), 51.4% male), and about half of the patients were male (statins: 51.4%; ezetimibe: 50.1%) (Table 1). Concomitant use of fibrates and liver dysfunction were more common in the ezetimibe group, whereas myocardial infarction (8.3% vs. 4.0%), stroke (13.8% vs. 10.3%), and heart failure (23.5% vs. 19.0%) were more frequently observed in the statin group. After inverse probability weighting, all covariates were balanced between the statin and ezetimibe groups, with aSMDs less than 0.10 (Table 1). The patient characteristics were balanced for all covariates between the lipophilic and hydrophilic statin subgroups, with aSMDs less than 0.10 before and after inverse probability weighting (eTable 5). In the fibrate groups, the median age was 56 years, and 72.0% of the patients were male (eTable 6).

**Table 1.**
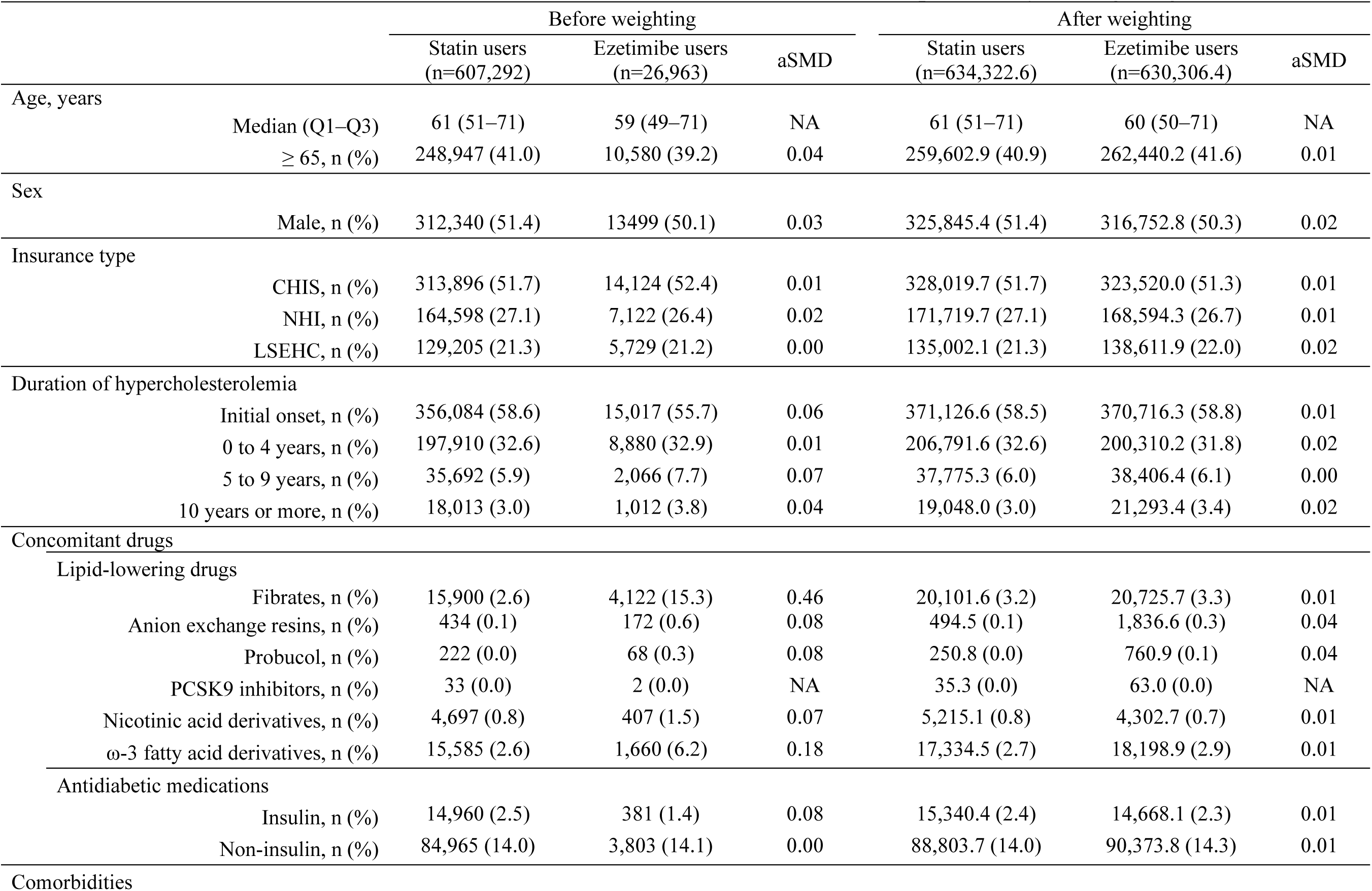
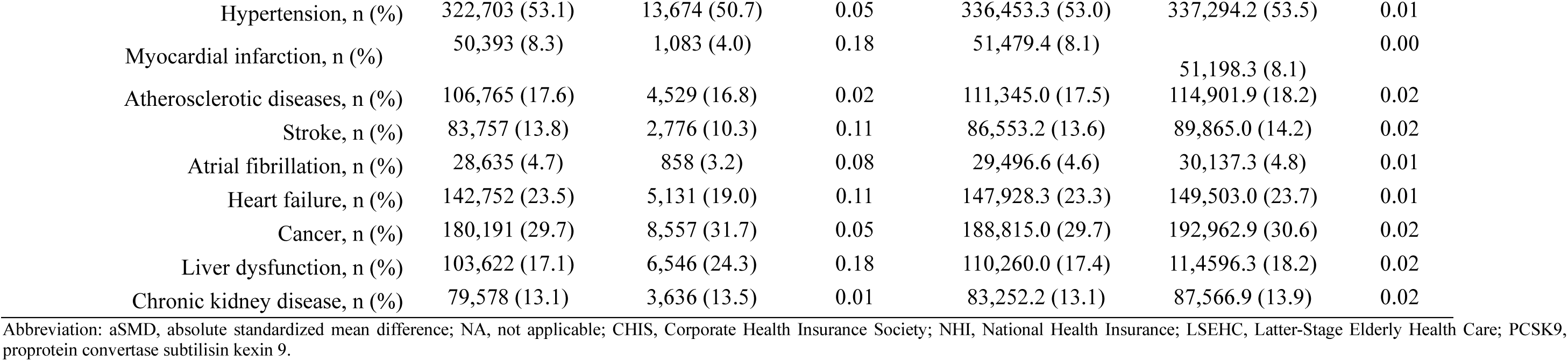
Patient characteristics of statin and ezetimibe users before and after inverse probability of weighting.

|  |  | Before weighting |  |  | After weighting |  |  |
| --- | --- | --- | --- | --- | --- | --- | --- |
|  |  | Statin users<br>(n=607,292) | Ezetimibe users<br>(n=26,963) | aSMD | Statin users<br>(n=634,322.6) | Ezetimibe users<br>(n=630,306.4) | aSMD |
| Age, years | Median (Q1–Q3) | 61 (51–71) | 59 (49–71) | NA | 61 (51–71) | 60 (50–71) | NA |
|  | ≥ 65, n (%) | 248,947 (41.0) | 10,580 (39.2) | 0.04 | 259,602.9 (40.9) | 262,440.2 (41.6) | 0.01 |
| Sex |  |  |  |  |  |  |  |
|  | Male, n (%) | 312,340 (51.4) | 13499 (50.1) | 0.03 | 325,845.4 (51.4) | 316,752.8 (50.3) | 0.02 |
| Insurance type |  |  |  |  |  |  |  |
|  | CHIS, n (%) | 313,896 (51.7) | 14,124 (52.4) | 0.01 | 328,019.7 (51.7) | 323,520.0 (51.3) | 0.01 |
|  | NHI, n (%) | 164,598 (27.1) | 7,122 (26.4) | 0.02 | 171,719.7 (27.1) | 168,594.3 (26.7) | 0.01 |
|  | LSEHC, n (%) | 129,205 (21.3) | 5,729 (21.2) | 0.00 | 135,002.1 (21.3) | 138,611.9 (22.0) | 0.02 |
| Duration of hypercholesterolemia |  |  |  |  |  |  |  |
|  | Initial onset, n (%) | 356,084 (58.6) | 15,017 (55.7) | 0.06 | 371,126.6 (58.5) | 370,716.3 (58.8) | 0.01 |
|  | 0 to 4 years, n (%) | 197,910 (32.6) | 8,880 (32.9) | 0.01 | 206,791.6 (32.6) | 200,310.2 (31.8) | 0.02 |
|  | 5 to 9 years, n (%) | 35,692 (5.9) | 2,066 (7.7) | 0.07 | 37,775.3 (6.0) | 38,406.4 (6.1) | 0.00 |
|  | 10 years or more, n (%) | 18,013 (3.0) | 1,012 (3.8) | 0.04 | 19,048.0 (3.0) | 21,293.4 (3.4) | 0.02 |
| Concomitant drugs |  |  |  |  |  |  |  |
| Lipid-lowering drugs |  |  |  |  |  |  |  |
|  | Fibrates, n (%) | 15,900 (2.6) | 4,122 (15.3) | 0.46 | 20,101.6 (3.2) | 20,725.7 (3.3) | 0.01 |
|  | Anion exchange resins, n (%) | 434 (0.1) | 172 (0.6) | 0.08 | 494.5 (0.1) | 1,836.6 (0.3) | 0.04 |
|  | Probucol, n (%) | 222 (0.0) | 68 (0.3) | 0.08 | 250.8 (0.0) | 760.9 (0.1) | 0.04 |
|  | PCSK9 inhibitors, n (%) | 33 (0.0) | 2 (0.0) | NA | 35.3 (0.0) | 63.0 (0.0) | NA |
|  | Nicotinic acid derivatives, n (%) | 4,697 (0.8) | 407 (1.5) | 0.07 | 5,215.1 (0.8) | 4,302.7 (0.7) | 0.01 |
|  | ω-3 fatty acid derivatives, n (%) | 15,585 (2.6) | 1,660 (6.2) | 0.18 | 17,334.5 (2.7) | 18,198.9 (2.9) | 0.01 |
| Antidiabetic medications |  |  |  |  |  |  |  |
|  | Insulin, n (%) | 14,960 (2.5) | 381 (1.4) | 0.08 | 15,340.4 (2.4) | 14,668.1 (2.3) | 0.01 |
|  | Non-insulin, n (%) | 84,965 (14.0) | 3,803 (14.1) | 0.00 | 88,803.7 (14.0) | 90,373.8 (14.3) | 0.01 |
| Comorbidities |  |  |  |  |  |  |  |

|  |  |  |  |  |  |  |
| --- | --- | --- | --- | --- | --- | --- |
| Hypertension, n (%) | 322,703 (53.1) | 13,674 (50.7) | 0.05 | 336,453.3 (53.0) | 337,294.2 (53.5) | 0.01 |
| Myocardial infarction, n (%) | 50,393 (8.3) | 1,083 (4.0) | 0.18 | 51,479.4 (8.1) | 51,198.3 (8.1) | 0.00 |
| Atherosclerotic diseases, n (%) | 106,765 (17.6) | 4,529 (16.8) | 0.02 | 111,345.0 (17.5) | 114,901.9 (18.2) | 0.02 |
| Stroke, n (%) | 83,757 (13.8) | 2,776 (10.3) | 0.11 | 86,553.2 (13.6) | 89,865.0 (14.2) | 0.02 |
| Atrial fibrillation, n (%) | 28,635 (4.7) | 858 (3.2) | 0.08 | 29,496.6 (4.6) | 30,137.3 (4.8) | 0.01 |
| Heart failure, n (%) | 142,752 (23.5) | 5,131 (19.0) | 0.11 | 147,928.3 (23.3) | 149,503.0 (23.7) | 0.01 |
| Cancer, n (%) | 180,191 (29.7) | 8,557 (31.7) | 0.05 | 188,815.0 (29.7) | 192,962.9 (30.6) | 0.02 |
| Liver dysfunction, n (%) | 103,622 (17.1) | 6,546 (24.3) | 0.18 | 110,260.0 (17.4) | 11,4596.3 (18.2) | 0.02 |
| Chronic kidney disease, n (%) | 79,578 (13.1) | 3,636 (13.5) | 0.01 | 83,252.2 (13.1) | 87,566.9 (13.9) | 0.02 |

Abbreviation: aSMD, absolute standardized mean difference; NA, not applicable; CHIS, Corporate Health Insurance Society; NHI, National Health Insurance; LSEHC, Latter-Stage Elderly Health Care; PCSK9, proprotein convertase subtilisin kexin 9.

The ezetimibe group had the highest incidence of ALS at 15.88 per 100,000 person-years (Table 2), followed by the statin group (6.82 per 100,000 person-years) and the fibrate group (4.28). The ALS incidence was 4.68 in the reference group consisting of all the members aged 40 years or older on the study database. The cumulative incidence in the statin group remained lower than the ezetimibe group throughout the follow-up period (Figure 2). By stratifying with lipophilicity of statins, the cumulative incidence in the lipophilic statin group stayed lower than the hydrophilic statin group.

**Figure 2.**
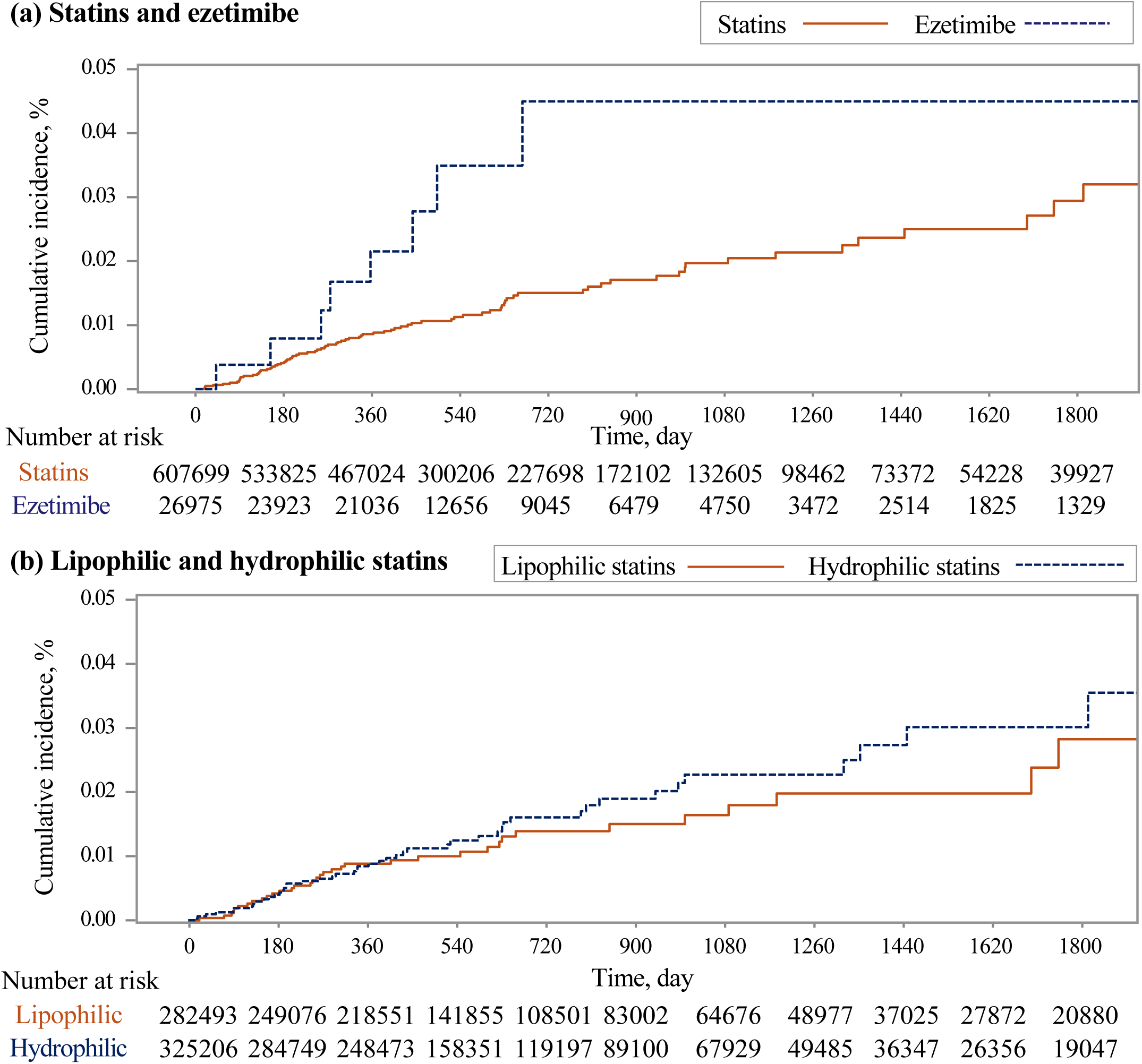
Parts a-b. Cumulative incidence curves of amyotrophic lateral sclerosis. (a) shows the cumulative incidence curves in the statin group (red solid line) and the ezetimibe group (blue dashed line). (b) Cumulative incidence curves in the lipophilic statin group (red solid line) and the hydrophilic statin group (blue dashed line).

**Table 2.** Incidence rate and hazard ratio of ALS.

| Group | N | Follow-up period (years) |  |  | Number of events | Incidence rate (/100,000 py) | Crude HR (95% CI) | Adjusted HR (95% CI) | P value |
| --- | --- | --- | --- | --- | --- | --- | --- | --- | --- |
|  |  | Median | Q1–Q3 | Total py |  |  |  |  |  |
| Statin | 607,699 | 1.46 | 1.03–2.71 | 1,217,819.34 | 83 | 6.82 | 0.44 (0.21–0.91) | 0.42 (0.19–0.92) | .03 |
| Ezetimibe | 26,975 | 1.39 | 1.03–2.40 | 50,381.30 | 8 | 15.88 | 1.00 (Reference) | 1.00 (Reference) | NA |
| Lipophilic statin | 282,493 | 1.48 | 1.03–2.79 | 581,651.63 | 36 | 6.19 | 0.85 (0.55–1.31) | 0.84 (0.55–1.31) | .46 |
| Hydrophilic statin | 325,206 | 1.44 | 1.02–2.64 | 636,167.71 | 47 | 7.39 | 1.00 (Reference) | 1.00 (Reference) | NA |
| Fibrate | 114,043 | 1.33 | 0.89–2.45 | 210,368.60 | 9 | 4.28 | NA | NA | NA |

Abbreviation: py, person-year; HR, hazard ratio; CI, confidence interval; NA, not applicable.

The mean LDL cholesterol levels prior to the index date were higher in the statin (162.8 ± 30.8 mg/dL) and ezetimibe (171.0 ± 28.6 mg/dL) groups compared to the fibrate group (111.8 ± 33.8 mg/dL, Figure 3). After treatment initiation, mean LDL cholesterol levels decreased to below 140 mg/dL in both groups. The statin group had a significantly lower hazard of ALS onset than the ezetimibe group (adjusted HR [95% CI]: 0.42 [0.19–0.92], p = 0.03) (Table 2). In all sensitivity analyses, the point estimates of the HRs consistently remained below 1 (eTable 7).

**Figure 3.**
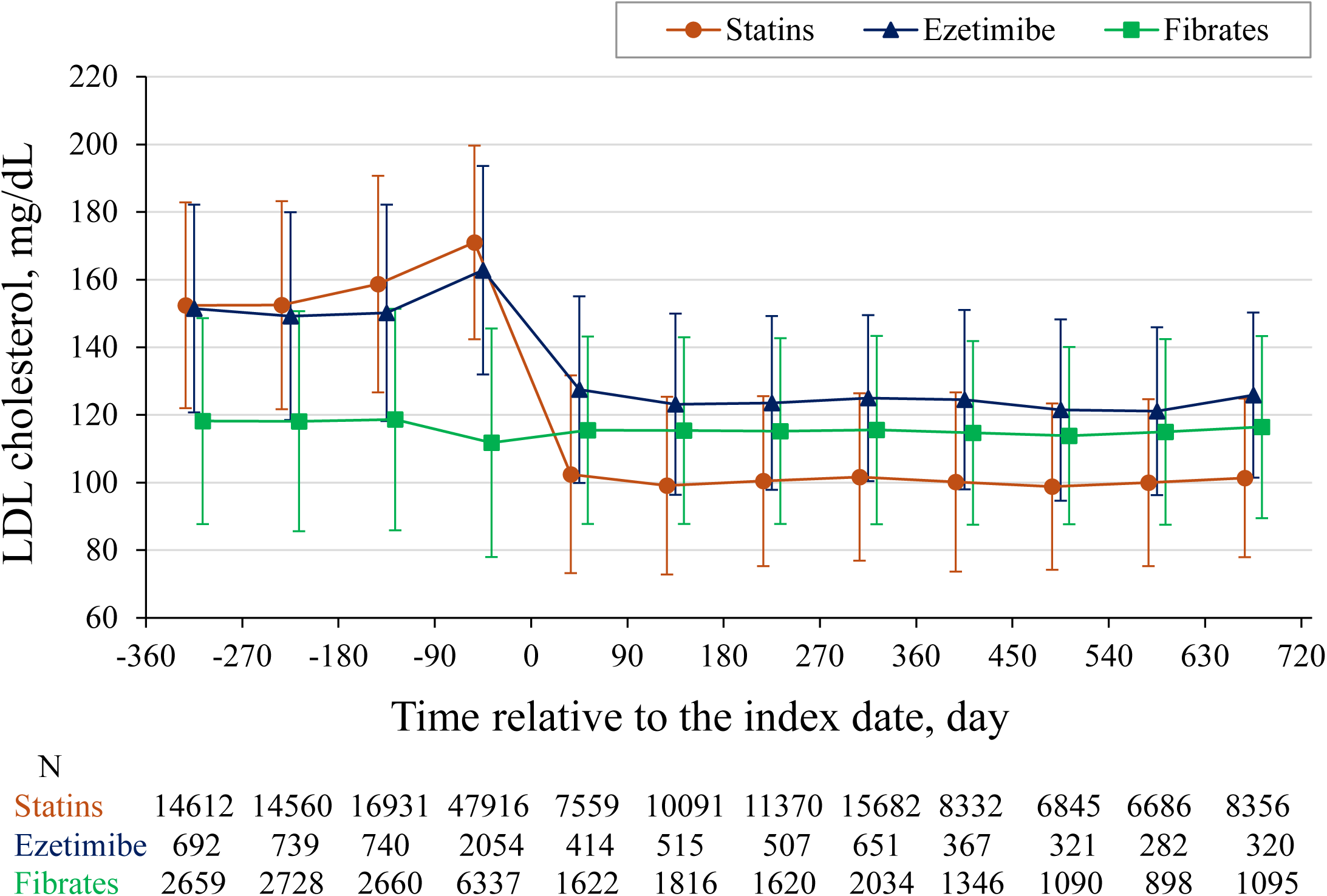
Time series data of low-density lipoprotein (LDL) cholesterol concentrations. Values and error bars represent the mean and standard deviation, respectively. Day 0 indicates the index date. LDL cholesterol levels were evaluated every 90 days from 360 days before to 720 days after Day 0. At each time point, patients with available LDL cholesterol measurements during the corresponding period were included in the analysis.

The adjusted HR for lipophilic statins versus hydrophilic statins was 0.84 (95% CI: 0.55–1.31). While the point estimate of the adjusted HR exceeded 1 in the sensitivity analysis with the stricter outcome definition (adjusted HR [95% CI]): 1.11 [0.53–2.32]), it consistently remained below 1 in all other sensitivity analyses (eTable 7).

### *In vitro* experimental study

Both ezetimibe and fenofibrate did not show neurotoxicity against human spinal motor neurons (eFigure 2).

## Discussion

This active-comparator, new-user cohort study of patients with hypercholesterolemia demonstrated that statin use was associated with a lower hazard of ALS onset when compared with ezetimibe use. In contrast, we did not observe a significant difference in ALS risk between lipophilic and hydrophilic statins. In our *in vitro* toxicity assays using human iPSC-derived spinal motor neurons, neither fenofibrate nor ezetimibe elicited apparent neurotoxic effects across the entire range of concentrations tested, from the lowest to the highest exposure levels.

Given that high LDL-cholesterol levels is one of the risk factors of ALS, and were similar between the statin group and the ezetimibe group at baseline of the present study, the finding of a lower hazard of ALS in the statin group suggests that statins mitigate the ALS risk through pathways not affected by ezetimibe. This interpretation is further supported by the finding that a lower incidence rate was observed in the fibrate group, which is likely to represent dyslipidemia patients with normal LDL-cholesterol levels. In addition, the incidence of ALS in statin users was only slightly higher than that of fibrate users. This modest difference between the statin group and the fibrate group is consistent with previous reports that have indicated there is either no difference, or a slightly higher risk of ALS, in statin users compared with nonusers.^16,17,20,21^ Our *in vitro* analyses indicated no neurotoxicity by ezetimibe, denying the possibility that ezetimibe would have a detrimental effect of CNS to increase the risk of ALS. Previous clinical studies may have been unable to detect a risk-reducing effect of statins because they compared statin users, who typically had elevated LDL cholesterol before treatment, with non-users who generally had normal LDL levels. This imbalance in baseline LDL cholesterol likely obscured any reduction in ALS risk attributable to statin therapy.^16,17,20,21^ If statins reduce the excess ALS risk associated with elevated LDL cholesterol, then statin users who begin treatment with high LDL levels may have an ALS risk similar to that of non-users that have normal LDL levels. As a result, prior studies would not detect a significant difference between the groups, even if statins were mitigating ALS risk.

This study has several notable strengths. First, we established two comparable groups of hypercholesterolemia patients to ensure comparison validity, with regard to baseline lipid status, by employing an active-comparator design. Second, the validity of our ALS case definition is supported by the fact that the incidence of ALS among adults aged 40 years or older in our databases was similar to previously reported national incidence estimates.^30^ Third, our iPS cells-based *in vitro* experiments further support our interpretation of the findings.

Given that both statins and ezetimibe lower circulating LDL cholesterol, the lower ALS risk observed with statins is unlikely to be explained solely by LDL reduction and may instead reflect unique mechanisms of action between the two drugs. The ability of statins to decrease ALS risk may involve inhibition of cholesterol biosynthesis in motor neurons, reduction of oxidative damage, and reduced activation of astrocytes.^31,32^ In addition, clinical and experimental studies of ropinirole have reported slowed ALS progression, potentially via suppression of the SREBP2–cholesterol synthesis pathway in the brain.^29^ In our *in vitro* analyses, using iPS cell-derived spinal motor neurons from familial ALS patients, two lipophilic statins atorvastatin and pitavastatin restored neurite outgrowth, which correlates with disease progression in ALS.^29^ Together, these findings suggest that the biological actions of statins on ALS onset may differ from those of ezetimibe.

Comparison among statin subclasses did not reveal any effect of lipophilicity on the risk of ALS. Although lipophilic statins may enter the central nervous system more readily because of their fat solubility, hydrophilic statins can still cross the blood–brain barrier via organic anion–transporting polypeptides.^33^ The subclass analysis was directionally consistent with the main findings, suggesting common class-effects of statins, although these comparisons were exploratory and not designed to detect modest differences between individual statins.

Our findings suggest that discontinuing or avoiding statins due to concern on the risk of ALS is likely not necessary. However, these findings should not be overgeneralized beyond patients with hypercholesterolemia, as the effect of statins on the ALS risk in the general population with normal LDL cholesterol levels remains unknown. Further, our study did not examine the effect of statins after ALS onset. Current evidence on whether statins modify prognosis in patients already diagnosed with ALS remains inconclusive, although most studies have reported no significant association.^34–37^

## Limitations

Although, to our knowledge, this is the first active-comparator, new-user cohort study to evaluate the effects of statin on ALS onset, it has several limitations. First, although this observational study adjusted for a wide range of confounding factors, the influence of potential unmeasured confounders, such as smoking history and body-mass index, cannot be ruled out. Second, since we defined the onset of ALS based on insurance claims, outcome misclassification is possible. Third, because the number of patients within each statin subclass was small, the statistical power to detect the different net effects on ALS incidence by statin subclasses may have been limited. Fourth, because the type of ALS was not available in the claims records, we could not assess whether the effect of statins differs by ALS type.

## Conclusion

Our findings indicate that statins may lower the risk of ALS onset among patients with hypercholesterolemia. As ezetimibe also lowers circulating LDL cholesterol, the lower ALS risk observed with statins is unlikely to be explained solely by LDL reduction, and may therefore reflect effects specific to statins beyond their lowering effect on circulating LDL cholesterol. Although subclass analyses were exploratory, the overall pattern was consistent across statins. These findings suggest that concern about ALS risk should not deter statin use in patients with hypercholesterolemia and point to the need for further research into lipid-related pathways in ALS.

## Data Availability

The data supporting the results of this study are obtained from JMDC Inc. and DeSC Healthcare, Inc. However, they are not publicly available and were used under license for the current study.

## Acknowledgement

We would like to thank Dr. Shinya Yamanaka, Dr. Keisuke Okita, and Dr. Makoto Nakagawa (CiRA, Kyoto University) for kindly providing 201B7, Dr. Scott Behie (Keio University) for editing the manuscript, and Ms. Shiho Nakamura and Ms. Fumiko Ozawa (KRM, Keio University) for their assistance with iPSC culture and data acquisition.

## Author contributions

Concept and design: Okada, Morimoto, Takahashi, Okano, Urushihara.

Acquisition, analysis, or interpretation of data: Okada, Morimoto, Takahashi, Okano, Urushihara.

Drafting of the manuscript: Okada, Morimoto.

Critical review of the manuscript for important intellectual content: Morimoto, Takahashi, Okano, Urushihara.

Statistical analysis: Okada

Administrative, technical, or material support: Urushihara.

Supervision: Okano, Urushihara.

## Information on author access to data

Okada and Urushihara had full access to all of the data in the study and take responsibility for the integrity of the data and accuracy of the data analysis.

