## Supplementary file for "Statin use and risk of amyotrophic lateral sclerosis: An active-comparator, new-user cohort study"

### List of contents

**eMethod 1. *In vitro* experimental study** 2
**eTable 1. Codelist of exposure drugs** 4
**eTable 2. Codelist of amyotrophic lateral sclerosis** 5
**eTable 3. Codelist of concomitant drugs** 6
**eTable 4. Codelist of comorbidities** 9
**eTable 5. Patient characteristics of statins, ezetimibe, and fibrate users before propensity score weighting** エラー! ブックマークが定義されていません。
**eTable 6. Patient characteristics of lipophilic and hydrophilic statin users after propensity score weighting** 10
**eTable 7. Adjusted hazard ratios based on the sensitivity analyses** 14
**eFigure 1. Design diagram** 15
**eFigure 2. Longitudinal effects of fenofibrate and ezetimibe on neurite dynamics in healthy iPSC-derived motor neurons** 16
**References** 18

### eMethod 1. *In vitro* experimental study

#### Research Ethics

All procedures involving human participants were performed in accordance with the principles of the Declaration of Helsinki and were approved by the Ethics Committee of Keio University School of Medicine (approval number 20080016).

#### Human Feeder-Free iPSC Culture

Human induced pluripotent stem cells (iPSCs) were cultured under feeder-free conditions in StemFit AK02N medium (Ajinomoto, Tokyo, Japan). For passage, cells were dissociated with 0.5 × TrypLE Select (Thermo Fisher Scientific, Waltham, MA, USA) and replated at a density of 0.3–1 × 10^4 cells per well onto six-well plates pre-coated with iMatrix-511 at 2 µL/mL (Matrixome, Osaka, Japan; Laminin 511E8, FUJIFILM Wako Pure Chemical Corp., Tokyo, Japan). The ROCK inhibitor Y27632 (10 µM; Nacalai, Kyoto, Japan) was added only during the first 24 hours after seeding. Culture medium was subsequently replaced every other day. The healthy control iPSC line 201B7 was used in this study.

#### Human spinal lower motor neuron (LMN) Induction from iPSCs Using Sendai Viruses

Lower motor neurons (LMNs) were generated from iPSCs through induction of a chemically transitional embryoid body-like state (CTraS) as described previously (1-3), followed by infection with SeV-LHX3-NGN2-ISL1 (Repli-tech Co., Ltd., Tokyo, Japan). In brief, iPSCs were plated in StemFit AK02N (Ajinomoto) supplemented with 10 µM Y27632 (Nacalai) and 2 µL/mL iMatrix-511 (Matrixome), and maintained for 5 days at 37°C in a humidified atmosphere containing 5% CO2. The medium was changed on the day after seeding to remove Y27632 and then renewed every 2 days.

To induce the EB-like CTraS state, the culture medium was switched to a chemical induction medium consisting of StemFit AK02N supplemented with 3 µM SB431542 (Sigma-Aldrich, St. Louis, MO, USA), 3 µM CHIR99021 (Cayman, Ann Arbor, MI, USA), and 3 µM dorsomorphin (Santa Cruz, Dallas, TX, USA) for 7 days (Fujimori et al., 2017). During this induction period, the medium was replaced daily.

After 7 days of chemical induction, iPSC colonies were dissociated into single cells using 0.5 × TrypLE Select and plated onto culture plates coated with 0.0001% poly-L-lysine (PLL; Sigma-Aldrich), 10 µL/mL Matrigel (Thermo Fisher Scientific), and 6.4 µL/mL iMatrix-511 (Matrixome). PLL was diluted in distilled water, applied to the wells, and incubated overnight at 37°C. Following removal of PLL, wells were treated with a mixture of Matrigel and iMatrix-511 prepared in phosphate-buffered saline (PBS) and incubated at 37°C for at least 1 hour. This coating solution was aspirated immediately before cell seeding. The same coating conditions were used for 96-well plates.

Motor neuron (MN) medium consisted of KBM Neural Stem Cell medium (KOHJIN BIO, Saitama, Japan) supplemented with 2% B27 (Thermo Fisher Scientific), penicillin (100 U/mL)–streptomycin (100 µg/mL) (Thermo Fisher Scientific), 200 µM ascorbic acid, 10 ng/mL brain-derived neurotrophic factor (R&D Systems, Minneapolis, MN, USA), 10 ng/mL glial cell-derived neurotrophic factor (Alomone Labs, Jerusalem, Israel), 10 µM DAPT (Sigma-Aldrich), and 10 µM Y27632. Sendai virus was applied at a multiplicity of infection (MOI) of 5. Y27632 was removed from the MN medium after 1 day. The culture medium was changed on days 1, 3, 4, 7, 10, and 13, and subsequently every 3 days. DAPT was included in the medium until day 7. To eliminate proliferating cells, 2 µM PD0332991 (Sigma-Aldrich) was added on days 4 and 7. When the composition of the medium was not altered, only half of the MN medium volume was replaced to minimize stress on LMNs. Cells were maintained at 37°C under 4% O2 and 5% CO2 until day 21 for neurite analyses.

**Time-course analysis of neurite outgrowth**

For longitudinal assessment of neurite extension in iPSC-derived motor neuron cultures, live MNs grown in multi-well plates were imaged using the BioStation CT live-cell imaging system (Nikon Instruments, Tokyo, Japan). For each well, images from five distinct fields were acquired with a 10× objective lens. Time-lapse imaging was initiated 21 days after the start of differentiation, with an imaging interval of 12 hours. Neuronal cells were identified based on EGFP fluorescence, and neurite structures were automatically traced and quantified using CL-Quant (Nikon Instruments), a machine learning-based image analysis platform. For each time point, neurite length per neuron was normalized to the corresponding value at day in vitro 3 (DIV3).

LMN cultures seeded in 96-well plates were treated with fenofibrate (1, 10, or 100 μM) or ezetimibe (1, 10, or 100 nM) from day 4 to day 10 in vitro. For negative controls, cells were exposed either to vehicle (DMSO at the corresponding final concentration) or left untreated. From day 4 to day 21, neurite outgrowth was monitored by time-lapse imaging using a BioStation CT incubator microscope (Nikon) to evaluate changes in neurite length.

### eTable 1. Codelist of exposure drugs

| Drug name | Classification | YJ code (first 7 digits) |
|---|---|---|
| Ezetimibe / Atorvastatin | Ezetimibe / Lipophilic statin | 2189101 |
| Ezetimibe / Rosuvastatin | Ezetimibe / Hydrophilic statin | 2189102 |
| Amlodipine Besylate / Atorvastatin Calcium | Lipophilic statin | 2190101 |
| | | 2190102 |
| | | 2190103 |
| | | 2190104 |
| Pravastatin Sodium | Hydrophilic statin | 2189010 |
| Simvastatin | Lipophilic statin | 2189011 |
| Fluvastatin Sodium | Lipophilic statin | 2189012 |
| Atorvastatin Calcium Hydrate | Lipophilic statin | 2189015 |
| Pitavastatin Calcium | Lipophilic statin | 2189016 |
| Rosuvastatin Calcium | Hydrophilic statin | 2189017 |
| Ezetimibe | Ezetimibe | 2183001 |
| Clinofibrate | Fibrate | 2183002 |
| Clofibrate | Fibrate | 2183005 |
| Bezafibrate | Fibrate | 2183006 |
| Fenofibrate | Fibrate | 2183007 |
| Pemafibrate | Fibrate | 2189018 |

### eTable 2. Codelist of amyotrophic lateral sclerosis

| Disease name | Standard disease code [a] |
| --- | --- |
| Familial amyotrophic lateral sclerosis | 8842326 |
| Progressive bulbar palsy | 3352010 |
| Sporadic amyotrophic lateral sclerosis | 8846130 |
| Amyotrophic lateral sclerosis | 3352007 |
| Motor neuron disease | 3352003 |

[a] The codes are coded by MEDIS standard disease master maintained by Medical Information System Development Center.

### eTable 3. Codelist of concomitant drugs

| Drug name | Classification | YJ code (first 7 digits) |
|---|---|---|
| Colestimide | Anion exchange resin | 2189014 |
| Cholestyramine | Anion exchange resin | 2189009 |
| Probucol | Probucol | 2189008 |
| Evolocumab（genetical recombination） | PCSK9 inhibitor | 2189401 |
| Nicomol | Nicotinic acid derivative | 2189004 |
| Niceritrol | Nicotinic acid derivative | 2189005 |
| Tocopherol nicotinate | Nicotinic acid derivative | 2190006 |
| Ethyl icosapentate | ω-3 fatty acid derivative | 2189021 |
| | | 3399004 |
| Ethyl omega-3 fatty acids | ω-3 fatty acid derivative | 2189019 |
| Insulin human (genetical recombination) | Insulin | 2492403 |
| | | 2492413 |
| Insulin | Insulin | 2492406 |
| Insulin lispro (genetical recombination) | Insulin | 2492414 |
| | | 2492422 |
| Insulin aspart (genetical recombination) | Insulin | 2492415 |
| | | 2492423 |
| Insulin glulisine (genetical recombination) | Insulin | 2492418 |
| Insulin degludec (genetical recombination) / insulin aspart (genetical recombination) | Insulin | 2492500 |
| Protamine zinc insulin | Insulin | 2492411 |
| Insulin glargine (genetical recombination) | Insulin | 2492416 |
| | | 2492420 |
| | | 2492421 |
| Insulin detemir (genetical recombination) | Insulin | 2492417 |
| Insulin degludec (genetical recombination) | Insulin | 2492419 |
| Insulin glargine (genetical recombination) / lixisenatide | Insulin / non-insulin | 3969501 |
| Insulin degludec (genetical recombination) / liraglutide (genetical recombination) | Insulin / non-insulin | 3969500 |
| Metformin hydrochloride | Non-insulin | 3962002 |
| Buformin hydrochloride | Non-insulin | 3962001 |
| Gliclopyramide | Non-insulin | 3961002 |
| Glibenclamide | Non-insulin | 3961003 |
| Chlorpropamide | Non-insulin | 3961004 |

| Drug name | Classification | YJ code (first 7 digits) |
|---|---|---|
| Tolbutamide | Non-insulin | 3961006 |
| Gliclazide | Non-insulin | 3961007 |
| Glimepiride | Non-insulin | 3961008 |
| Acetohexamide | Non-insulin | 3961001 |
| Teneligliptin hydrobromide hydrate / canagliflozin hydrate | Non-insulin | 3969106 |
| Anagliptin / metformin hydrochloride | Non-insulin | 3969109 |
| Mitiglinide calcium hydrate / voglibose | Non-insulin | 3969102 |
| Sitagliptin phosphate hydrate / ipragliflozin L-proline | Non-insulin | 3969107 |
| Pioglitazone hydrochloride / metformin hydrochloride | Non-insulin | 3969100 |
| Pioglitazone hydrochloride / glimepiride | Non-insulin | 3969101 |
| Vildagliptin / metformin hydrochloride | Non-insulin | 3969104 |
| Alogliptin benzoate / pioglitazone hydrochloride | Non-insulin | 3969103 |
| Alogliptin benzoate / metformin hydrochloride | Non-insulin | 3969105 |
| Empagliflozin / linagliptin | Non-insulin | 3969108 |
| Acarbose | Non-insulin | 3969003 |
| Miglitol | Non-insulin | 3969009 |
| Voglibose | Non-insulin | 3969004 |
| Pioglitazone hydrochloride | Non-insulin | 3969007 |
| Trelagliptin succinate | Non-insulin | 3969024 |
| Omarigliptin | Non-insulin | 3969025 |
| Anagliptin | Non-insulin | 3969016 |
| Sitagliptin phosphate hydrate | Non-insulin | 3969010 |
| Vildagliptin | Non-insulin | 3969011 |
| Saxagliptin hydrate | Non-insulin | 3969017 |
| Alogliptin benzoate | Non-insulin | 3969012 |
| Linagliptin | Non-insulin | 3969014 |
| Teneligliptin hydrobromide hydrate | Non-insulin | 3969015 |
| Exenatide | Non-insulin | 2499411 |
| Liraglutide (genetical recombination) | Non-insulin | 2499410 |
| Lixisenatide | Non-insulin | 2499415 |
| Dulaglutide (genetical recombination) | Non-insulin | 2499416 |
| Semaglutide (genetical recombination) | Non-insulin | 2499014 |
| | | 2499418 |
| Tofogliflozin hydrate | Non-insulin | 3969021 |
| Dapagliflozin propylene glycol hydrate | Non-insulin | 3969019 |

| Drug name | Classification | YJ code (first 7 digits) |
|---|---|---|
| Canagliflozin hydrate | Non-insulin | 3969022 |
| Empagliflozin | Non-insulin | 3969023 |
| Ipragliflozin L-proline | Non-insulin | 3969018 |
| Luseogliflozin hydrate | Non-insulin | 3969020 |
| Repaglinide | Non-insulin | 3969013 |
| Nateglinide | Non-insulin | 3969006 |
| Mitiglinide calcium | Non-insulin | 3969008 |
| Imeglimin hydrochloride | Non-insulin | 3969026 |
| Tirzepatide | Non-insulin | 2499422 |

Abbreviation: PCSK9, proprotein convertase subtilisin kexin 9.

**eTable 4. Codelist of comorbidities**

| Disease name | ICD-10 code |
|---|---|
| Hypertension | I10–I15 |
| Myocardial infarction | I21–I23 |
| Atherosclerosis | I70–I74 |
| Stroke | I71–I79 |
| Atrial fibrillation | I48 |
| Heart failure | I50 |
| Cancer | C00–C96 |
| Liver disease | B150 |
| | B160 |
| | B162 |
| | B18 |
| | B190 |
| | I85 |
| | K700–K704 |
| | K709 |
| | K71–K74 |
| | K760 |
| | K766 |
| Chronic kidney disease | E102 |
| | E112 |
| | E142 |
| | N03 |
| | N05 |
| | N110 |
| | N14 |
| | N16 |
| | N18–N19 |
| | N269 |
| | Q611–Q614 |

**eTable 5. Patient characteristics of lipophilic and hydrophilic statin users before and after propensity score weighting**

| | Before weighting | | | After weighting | | |
|---|---|---|---|---|---|---|
| | Lipophilic statins (n=282,493) | Hydrophilic statins (n=325,206) | aSMD | Lipophilic statins (n= 634,322.6) | Hydrophilic statins (n= 630,306.4) | aSMD |
| Age, years | | | | | | |
| Median (Q1–Q3) | 61 (51–71) | 60 (51–71) | NA [a] | 61 (51–71) | 61 (51–71) | NA [a] |
| ≥ 65, n (%) | 116,959 (41.4) | 131,988 (40.6) | 0.02 | 249,027.6 (41.0) | 249,004.7 (41.0) | 0.00 |
| Sex | | | | | | |
| Male, n (%) | 144,111 (51.0) | 168,421 (51.8) | 0.02 | 312,435.5 (51.4) | 312,452.6 (51.4) | 0.00 |
| Insurance type | | | | | | |
| CHIS, n (%) | 144,694 (51.2) | 169,202 (52.0) | 0.02 | 313,832.8 (51.6) | 313,825.1 (51.6) | 0.00 |
| NHI, n (%) | 76,853 (27.2) | 87,745 (27.0) | 0.00 | 164,634.9 (27.1) | 164,629.7 (27.1) | 0.00 |
| LSEHC, n (%) | 60,946 (21.6) | 68,259 (21.0) | 0.01 | 129,249.7 (21.3) | 129,233.6 (21.3) | 0.00 |
| Duration of hypercholesterolemia | | | | | | |
| Initial onset, n (%) | 167,224 (59.2) | 188,860 (58.1) | 0.02 | 356,044.6 (58.6) | 356,033.1 (58.6) | 0.00 |
| 0 to 4 years, n (%) | 89,815 (31.8) | 108,095 (33.2) | 0.03 | 197,931.4 (32.6) | 197,923.4 (32.6) | 0.00 |
| 5 to 9 years, n (%) | 16,730 (5.9) | 18,962 (5.8) | 0.00 | 35,727.4 (5.9) | 35,716.6 (5.9) | 0.00 |
| 10 years or more, n (%) | 8,724 (3.1) | 9,289 (2.9) | 0.01 | 18,014.0 (3.0) | 18,015.4 (3.0) | 0.00 |
| Concomitant drugs | | | | | | |
| Lipid-lowering drugs | | | | | | |
| Fibrates, n (%) | 7,458 (2.6) | 8,442 (2.6) | 0.00 | 15,907.7 (2.6) | 15,907.2 (2.6) | 0.00 |
| Anion exchange resins, n (%) | 229 (0.1) | 205 (0.1) | 0.00 | 486.4 (0.1) | 391.6 (0.1) | 0.00 |
| Probucol, n (%) | 99 (0.0) | 123 (0.0) | NA [a] | 206.4 (0.0) | 236.5 (0.0) | NA [a] |
| PCSK9 inhibitors, n (%) | 14 (0.0) | 19 (0.0) | NA [a] | 30.7 (0.0) | 34.2 (0.0) | NA [a] |

| | | | | | | |
|---|---|---|---|---|---|---|
| Nicotinic acid derivatives, n (%) | 2,227 (0.8) | 2,470 (0.8) | 0.00 | 4,658.7 (0.8) | 4,743.7 (0.8) | 0.00 |
| ω-3 fatty acid derivatives, n (%) | 7,448 (2.6) | 8,137 (2.5) | 0.01 | 15,594.9 (2.6) | 15,582.5 (2.6) | 0.00 |
| Antidiabetic medications | | | | | | |
| Insulin, n (%) | 6,866 (2.4) | 8,094 (2.5) | 0.01 | 14,966.3 (2.5) | 14,962.5 (2.5) | 0.00 |
| Non-insulin, n (%) | 40,080 (14.2) | 44,885 (13.8) | 0.01 | 84,976.1 (14.0) | 84,970.8 (14.0) | 0.00 |
| Comorbidities | | | | | | |
| Hypertension, n (%) | 154,464 (54.7) | 168,239 (51.7) | 0.06 | 322,583.4 (53.1) | 322,622.7 (53.1) | 0.00 |
| Myocardial infarction, n (%) | 20,580 (7.3) | 29,813 (9.2) | 0.07 | 50,465.3 (8.3) | 50,426.6 (8.3) | 0.00 |
| Atherosclerotic diseases, n (%) | 49,728 (17.6) | 57,037 (17.5) | 0.00 | 106,744.6 (17.6) | 106,745.0 (17.6) | 0.00 |
| Stroke, n (%) | 39,060 (13.8) | 44,697 (13.7) | 0.00 | 83,899.8 (13.8) | 83,853.5 (13.8) | 0.00 |
| Atrial fibrillation, n (%) | 12,945 (4.6) | 15,690 (4.8) | 0.01 | 28,644.6 (4.7) | 28,640.1 (4.7) | 0.00 |
| Heart failure, n (%) | 64,259 (22.7) | 78,493 (24.1) | 0.03 | 142,803.4 (23.5) | 142,769.1 (23.5) | 0.00 |
| Cancer, n (%) | 83,800 (29.7) | 96,391 (29.6) | 0.00 | 180,256.3 (29.7) | 180,229.0 (29.7) | 0.00 |
| Liver dysfunction, n (%) | 48,192 (17.1) | 55,430 (17.0) | 0.00 | 103,654.9 (17.1) | 103,642.6 (17.1) | 0.00 |
| Chronic kidney disease, n (%) | 39,166 (13.9) | 40,412 (12.4) | 0.04 | 79,570.9 (13.1) | 79,575.2 (13.1) | 0.00 |

Abbreviation: aSMD, absolute standardized mean difference; NA, not applicable; CHIS, Corporate Health Insurance Society; NHI, National Health Insurance; LSEHC, Latter-Stage Elderly Health Care; PCSK9, proprotein convertase subtilisin kexin 9.

[a] The aSMD cannot be calculated.

**eTable 6. Patient characteristics of fibrates, statins, and ezetimibe users before propensity score weighting**

| | Fibrates (n=114,871) | Statins (n=607,292) | Ezetimibe (n=26,963) |
|---|---|---|---|
| Age, years | | | |
| Median (Q1–Q3) | 56 (46-68) | 61 (51–71) | 59 (49–71) |
| ≥ 65, n (%) | 36,389 (31.7) | 248,947 (41.0) | 10573 (39.2) |
| Sex | | | |
| Male, n (%) | 82,762 (72.0) | 312,340 (51.4) | 13499 (50.1) |
| Insurance type | | | |
| CHIS, n (%) | 64,648 (56.2) | 313,896 (51.7) | 14121 (52.4) |
| NHI, n (%) | 31,974 (27.8) | 164,598 (27.1) | 7115 (26.4) |
| LSEHC, n (%A) | 18,321 (15.9) | 129,205 (21.3) | 5727 (21.2) |
| Duration of hypercholesterolemia [a] | | | |
| Initial onset, n (%) | 41,700 (36.3) | 356,084 (58.6) | 15010 (55.7) |
| 0 to 4 years, n (%) | 52,786 (45.9) | 197,910 (32.6) | 8875 (32.9) |
| 5 to 9 years, n (%) | 13,330 (11.6) | 35,692 (5.9) | 2066 (7.7) |
| 10 years or more, n (%) | 7,127 (6.2) | 18,013 (3.0) | 1012 (3.8) |
| Concomitant drugs | | | |
| Lipid-lowering drugs | | | |
| Fibrates, n (%) | NA | 15,900 (2.6) | 4121 (15.3) |
| Anion exchange resins, n (%) | 191 (0.2) | 434 (0.1) | 172 (0.6) |
| Probucol, n (%) | 78 (0.1) | 222 (0.0) | 68 (0.3) |
| PCSK9 inhibitors, n (%) | 7 (0.0) | 33 (0.0) | 2 (0.0) |
| Nicotinic acid derivatives, n (%) | 1,519 (1.3) | 4,697 (0.8) | 407 (1.5) |
| ω-3 fatty acid derivatives, n (%) | 4,850 (4.2) | 15,585 (2.6) | 1660 (6.2) |
| Antidiabetic medications | | | |
| Insulin, n (%) | 1,932 (1.7) | 14,960 (2.5) | 381 (1.4) |
| Non-insulin, n (%) | 20,327 (17.7) | 84,965 (14.0) | 3799 (14.1) |
| Comorbidities | | | |
| Hypertension, n (%) | 63,459 (55.2) | 322,703 (53.1) | 13664 (50.7) |
| Myocardial infarction, n (%) | 3,635 (3.2) | 50,393 (8.3) | 1082 (4.0) |
| Atherosclerotic diseases, n (%) | 13,975 (12.2) | 106,765 (17.6) | 4528 (16.8) |
| Stroke, n (%) | 9,146 (8.0) | 83,757 (13.8) | 2775 (10.3) |
| Atrial fibrillation, n (%) | 3,428 (3.0) | 28,635 (4.7) | 857 (3.2) |
| Heart failure, n (%) | 18,762 (16.3) | 142,752 (23.5) | 5127 (19.0) |
| Cancer, n (%) | 33,571 (29.2) | 180,191 (29.7) | 8554 (31.7) |
| Liver dysfunction, n (%) | 33,381 (29.0) | 103,622 (17.1) | 6540 (24.3) |

| | | | |
|---|---|---|---|
| Chronic kidney disease, n (%) | 14,330 (12.5) | 79,578 (13.1) | 3635 (13.5) |

Abbreviation: NA, not applicable; CHIS, Corporate Health Insurance Society; NHI, National Health Insurance; LSEHC, Latter-Stage Elderly Health Care; PCSK9, proprotein convertase subtilisin kexin 9.

[a] For fibrate users, the results for the duration of dyslipidemia are presented.

### eTable 7. Adjusted hazard ratios based on the sensitivity analyses

| Analysis condition | | Adjusted HR (95% CI) |
|---|---|---|
| **Statins vs. ezetimibe** | | |
| Main analysis | | 0.42 (0.19–0.92) |
| Sensitivity analysis | Stricter outcome definition [a] | 0.47 (0.11–1.92) |
| | Longer follow-up after treatment discontinuation or switching [b] | 0.44 (0.21–0.92) |
| | Shorter follow-up after treatment discontinuation or switching [c] | 0.41 (0.18–0.97) |
| | 60-day gap period instead of 100 days | 0.40 (0.18–0.88) |
| **Lipophilic statins vs. hydrophilic statins** | | |
| Main analysis | | 0.84 (0.55–1.31) |
| Sensitivity analysis | Stricter outcome definition | 1.11 (0.53–2.32) |
| | Longer follow-up after treatment discontinuation or switching [b] | 0.89 (0.59–1.34) |
| | Shorter follow-up after treatment discontinuation or switching [c] | 0.77 (0.48–1.22) |
| | 60-day gap period instead of 100 days | 0.88 (0.57–1.37) |

Abbreviation: HR, hazard ratio; CI, confidence interval.

[a] An outcome definition for ALS that included, in addition to diagnostic record of ALS, the presence of a prescription record for an ALS treatment (riluzole or edaravone) in the same month as the diagnostic record.

[b] Replacing the 365-day window after treatment discontinuation or switching with 730 days.

[c] Replacing the 365-day window after treatment discontinuation or switching with 180 days.

[d] In this analysis condition, no events occurred in the standard statin group, and therefore HR estimates could not be obtained from the Cox proportional hazards model.

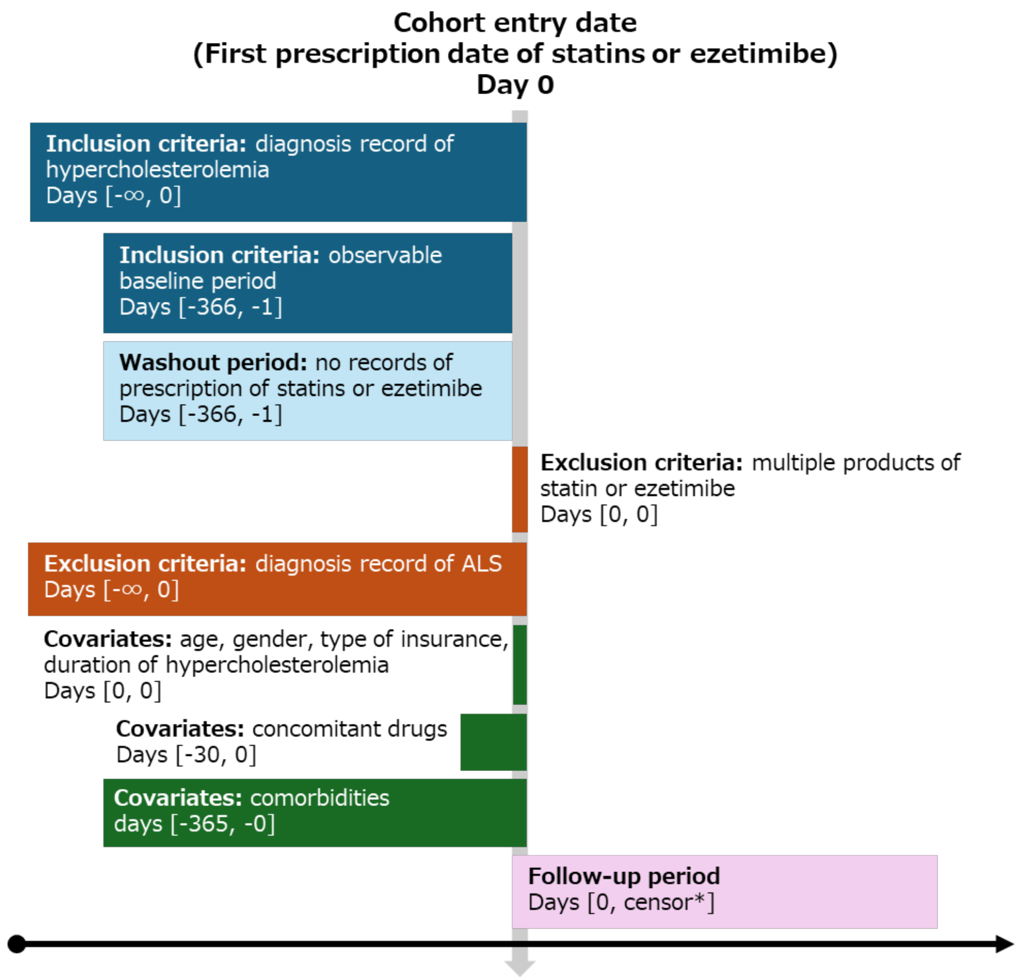


### eFigure 1. Design diagram

ALS, amyotrophic lateral sclerosis.

* The earliest of the following dates: 365 days after treatment discontinuation or switching, date of ALS onset, end of the observable data in the database.

(A)

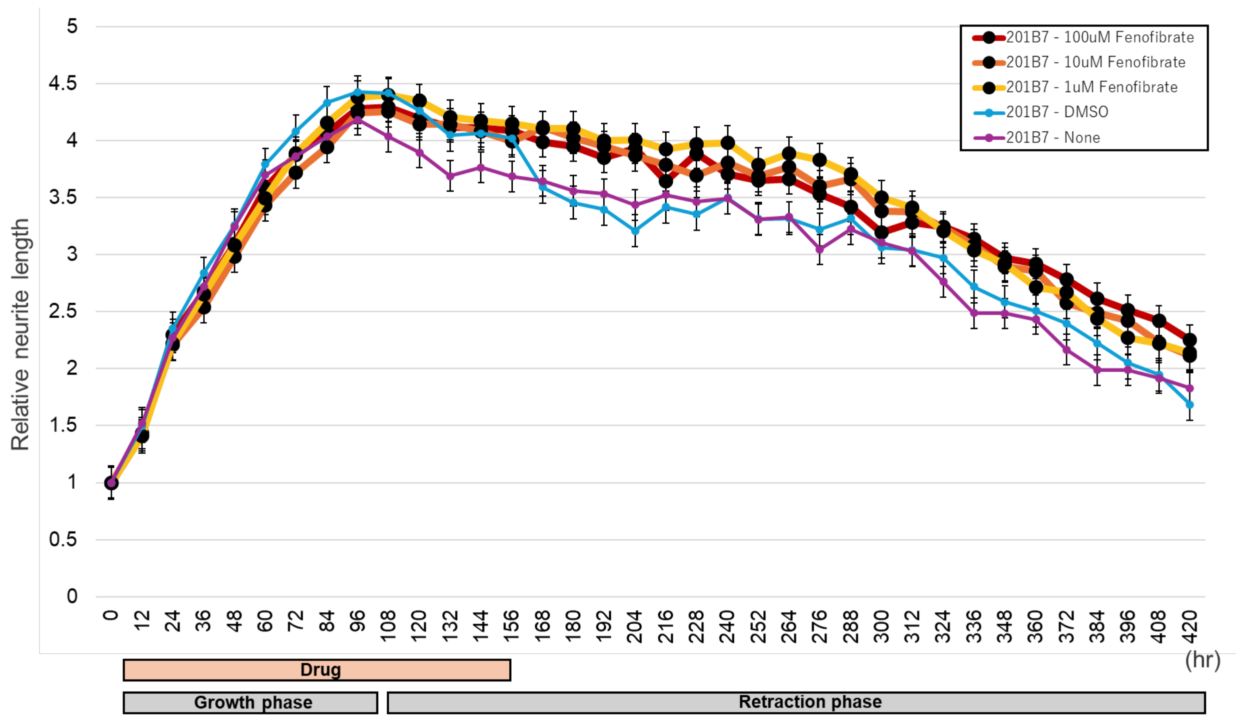


(B)

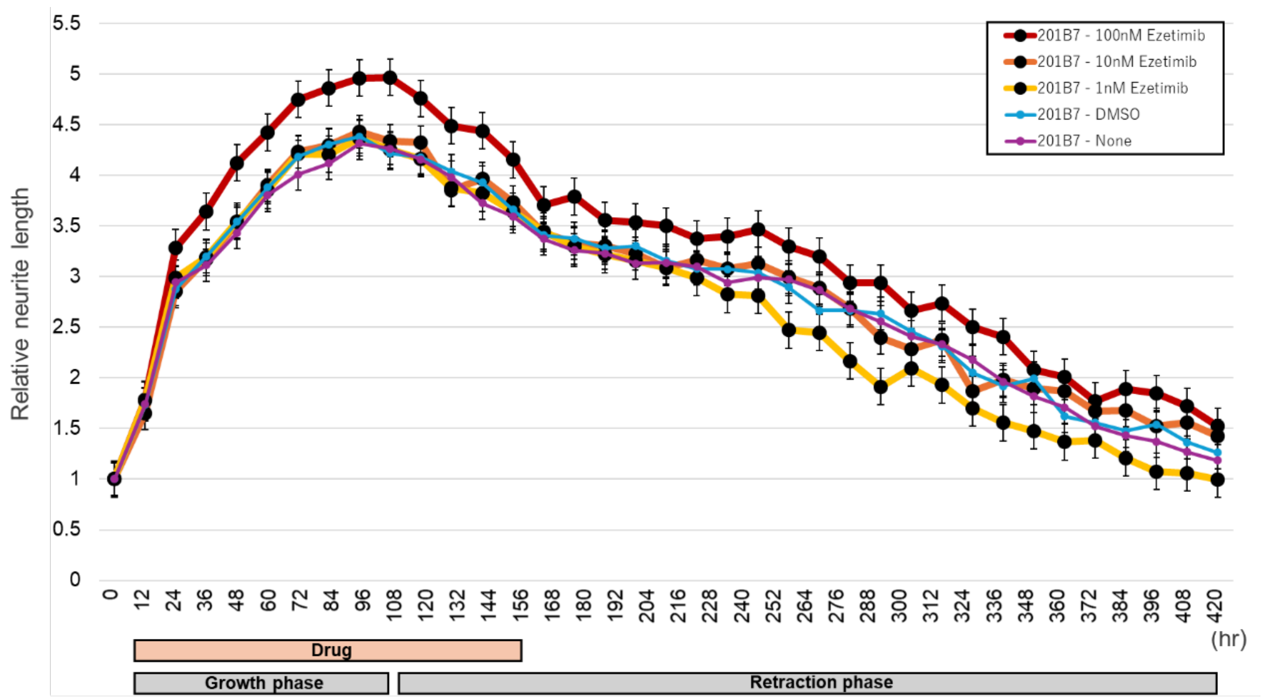


**eFigure 2. Longitudinal effects of fenofibrate and ezetimibe on neurite dynamics in healthy iPSC-derived motor neurons**

(A) Time-course of normalized neurite length in 201B7 iPSC-derived lower motor neurons treated with fenofibrate (1, 10, or 100 μM) from day 4 to day 10 *in vitro*, compared with DMSO vehicle and untreated controls. (B) Time-course of normalized neurite length in 201B7 motor neurons treated with ezetimibe (1, 10, or 100 nM) over the same period. Neurons were cultured in 96-well plates and imaged using a BioStation CT live-cell imaging system with a 10× objective every 12 hr from day in vitro (DIV) 3 to DIV21. Neurites were identified based on EGFP fluorescence and automatically traced and quantified with CL-Quant software. Neurite length per neuron at each time point was normalized to the mean value at DIV3 (time 0). The pink bar below the x-axis indicates the drug treatment period (day 4–10), and gray bars denote the neurite growth and retraction phases. Data points represent the mean relative neurite length from three technical replicates (n = 3 wells, 5 fields per well; 15 measurements per time point), and error bars indicate the standard error of the mean (SE).
